# Predicting Carbapenem Resistance in Hospitalized Patients Using Machine Learning: A Retrospective Analysis of the MIMIC-III Database

**DOI:** 10.1101/2025.11.24.25340702

**Authors:** Addiel Ulises de Alba Solis, Ivette Sarahi Ocampo Morales, Eduardo Gómez Sánchez, Hugo Enrique Chávez Chávez

## Abstract

Carbapenem-resistant Gram-negative bacteria (CR-GNB) represent a major health challenge due to limited therapeutic options, increased morbidity, and extended hospital stays (Ham et al., 2021). Early prediction of carbapenem resistance can optimize empirical antibiotic therapy and reduce unnecessary use of broad-spectrum antibiotics. This study developed and validated an artificial intelligence model to predict carbapenem resistance among hospitalized and ICU patients using clinical, demographic, laboratory, and antibiogram data. A retrospective analysis was performed on 94,000 hospitalized patients at Beth Israel Deaconess Medical Centre, utilizing the MIMIC-III (v1.4) database (Johnson et al., 2016). Predictor variables included were demographic characteristics, comorbidities, ICU admission, antibiotic exposure history, inflammatory and biochemical markers, and antibiotic susceptibility test results. Three artificial intelligence algorithms were evaluated: decision trees, random forests, and XGBoost. Model performance was assessed using AUC, precision, sensitivity, specificity, and confusion matrices. The XGBoost model demonstrated the highest performance, achieving an AUC of 0.95, precision of 0.98, sensitivity of 0.90, and specificity of 0.99. These results demonstrate strong discrimination ability and significant potential for integration into clinical workflows. The findings support the use of machine learning to enhance infection prevention, improve antibiotic stewardship, and inform early clinical decision-making.

**METHODS:** A retrospective study was performed using clinical and antibiotic-susceptibility test reports from hospitalized patients with confirmed bacterial infections, drawn from a cohort of 94,000 patients admitted to Beth Israel Deaconess Medical Centre in Boston, Massachusetts, USA, utilizing the MIMIC-III (v1.4) database. Three artificial intelligence algorithms were assessed: decision trees, random forests, and XGBoost. Data preprocessing ensured quality and completeness, followed by division into training and validation sets. Model performance was evaluated using accuracy, sensitivity, specificity, and confusion matrices. The study required access to electronic health records, computational servers for data processing and storage, specialized Python machine learning code, and a coding platform for prototyping on a multicore virtual machine. Python, along with libraries such as pandas, NumPy, scikit-learn, matplotlib, and seaborn, was used, primarily leveraging the collaborative development environment Google Colab.

**RESULTS:** Three models were trained using clinical, laboratory, and microbiological data from 94,000 hospitalized patients, with a subset of 3,277 patients with carbapenem-resistant infections. The analysis focused on patients with confirmed bacterial infections and antibiotic resistance, particularly within the ESKAPE group (Enterococcus faecium, *S*taphylococcus aureus, *K*lebsiella pneumoniae, *A*cinetobacter baumannii, *P*seudomonas aeruginosa, and *E*nterobacter species), which cause most hospital-acquired infections (McGuire et al., 2021). The most favorable results were obtained using the XGBoost model, yielding:

- **AUC:** 0.955
- **Precision:** 0.989
- **Sensitivity:** 0.909
- **Specificity:** 0.993

The model demonstrated high reliability in predicting carbapenem before culture results were available, utilizing data accessible within the first hours of ICU admission. Early prediction enabled simulation of clinical scenarios in which the model recommended more timely antibiotic treatments, potentially reducing unnecessary exposure to broad-spectrum antibiotics when appropriately applied.

**CONCLUSION:** The predictive model demonstrated high accuracy and real-world clinical applicability for anticipating carbapenem resistance in critically ill patients, including, in some cases, septic patients. Its use in the first hours of hospitalization could:

- Guide the initial antibiotic selection more appropriately.
- Reduce unnecessary carbapenem use to help contain antimicrobial resistance.
- Decrease the time to effective treatment to improve clinical outcomes.
- Optimize ICU resources by reducing complications associated with inadequate treatments.

In clinical practice, this tool could be integrated into electronic health systems to automatically alert medical staff to a high risk of resistance, thereby streamlining critical decision-making in high-demand environments such as the ICU.

## Introduction

Antimicrobial resistance (AMR) is widely recognized as one of the most urgent public health threats in current medical practice (Ham et al., 2021). Among resistant organisms, carbapenem-resistant Gram-negative bacteria pose a particular danger because they often affect critically ill patients and are associated with high mortality. The resistance mechanisms of these pathogens, including carbapenemase production, efflux pump overexpression, and porin loss confer broad protection against β-lactam antibiotics, leaving physicians with limited treatment options (Raman et al., 2018).

The prevalence of CR-GNB has increased the complexity of managing infections in intensive care units and higher specialized hospitals (X. Li et al., 2024). These microorganisms disproportionately affect patients with prolonged hospital stays, invasive catheter contamination, correlated immunosuppression, or extensive prior antibiotic exposure (Salomão et al., 2020). Consequently, delayed recognition of carbapenem resistance can lead to inappropriate empiric therapy, septic deterioration, and extended hospitalization (Zilberberg et al., 2017). Optimizing and correctly choosing initial therapy remains a crucial step in reducing mortality and preventing the further spread of resistant strains and their mutation rates.

Machine learning and artificial intelligence have emerged as promising methods for predicting patient outcomes using large, multivariable clinical datasets such as the MIMIC-III database (Li et al., 2025). Unlike traditional statistical models, machine learning algorithms such as XGBoost can detect nonlinear relationships, variable interactions, and non-documented patterns that may be difficult for health professionals to correlate and interpret (Enguehard et al., 2022). Prior research has demonstrated that these types of models can improve the prediction of sepsis, hospital mortality, and adverse drug reactions, but their application to antimicrobial resistance prediction remains relatively underexplored (Rosnati et al., 2019).

The MIMIC-III database offers a comprehensive source of anonymized clinical data, including laboratory results, vital signs, demographics, ICD diagnoses, medication administration records, and microbiology reports (Kim & J., 2024). This dataset supports the development of predictive models based on real-world clinical data. In this study, the dataset was used to construct a machine learning model capable of predicting carbapenem resistance at the point of clinical suspicion, before culture and susceptibility results are available. The objective is to assist clinicians in selecting more precise empiric antibiotic therapy, thereby reducing unnecessary carbapenem use and improving patient outcomes.

This manuscript details the methodology, development, validation, and clinical relevance of an artificial intelligence model designed to predict carbapenem resistance among hospitalized and ICU-admitted patients.

## Materials and Methods

### Study Design, Data Source, and Inclusion Criteria

We conducted a retrospective observational study using the MIMIC-III v1.4 dataset, a comprehensive, curated database of de-identified health records from hospitalized patients, including ICU patients at the Beth Israel Deaconess Medical Centre in Boston, Massachusetts, USA, from 2001 to 2012. The study population included all adult patients with available microbiological cultures and positive carbapenem-susceptibility test results. After applying inclusion and exclusion criteria, 3,277 unique admissions were included. To narrow the target population, the criteria stipulated that patients should have available antibiotic resistance testing results showing Carbapenem-resistant Gram-negative bacteria and that they had had any fluid or blood samples collected at admission or during hospitalization. This filtering and inclusion criteria yielded a sample of 3,277 patients who tested positive for carbapenem resistance. Image 1 illustrates the study’s design and data sources, modelling effort, and the future application of the software solution.

**Image 1.**
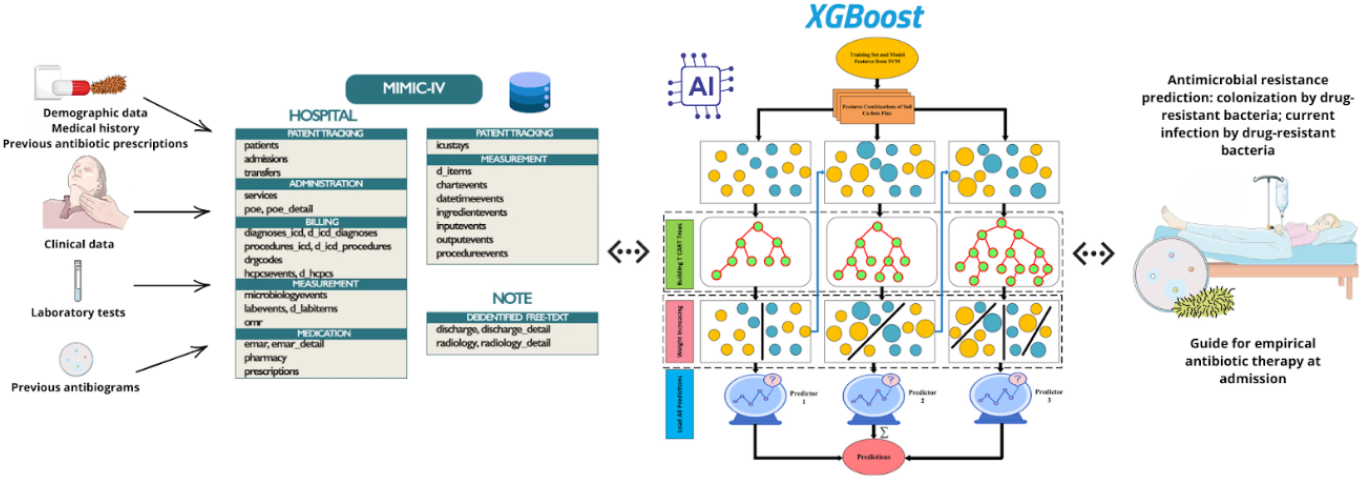
Study and prototype design of the flow of information and machine learning modelling applied to a clinical setting.

**Image 2.**
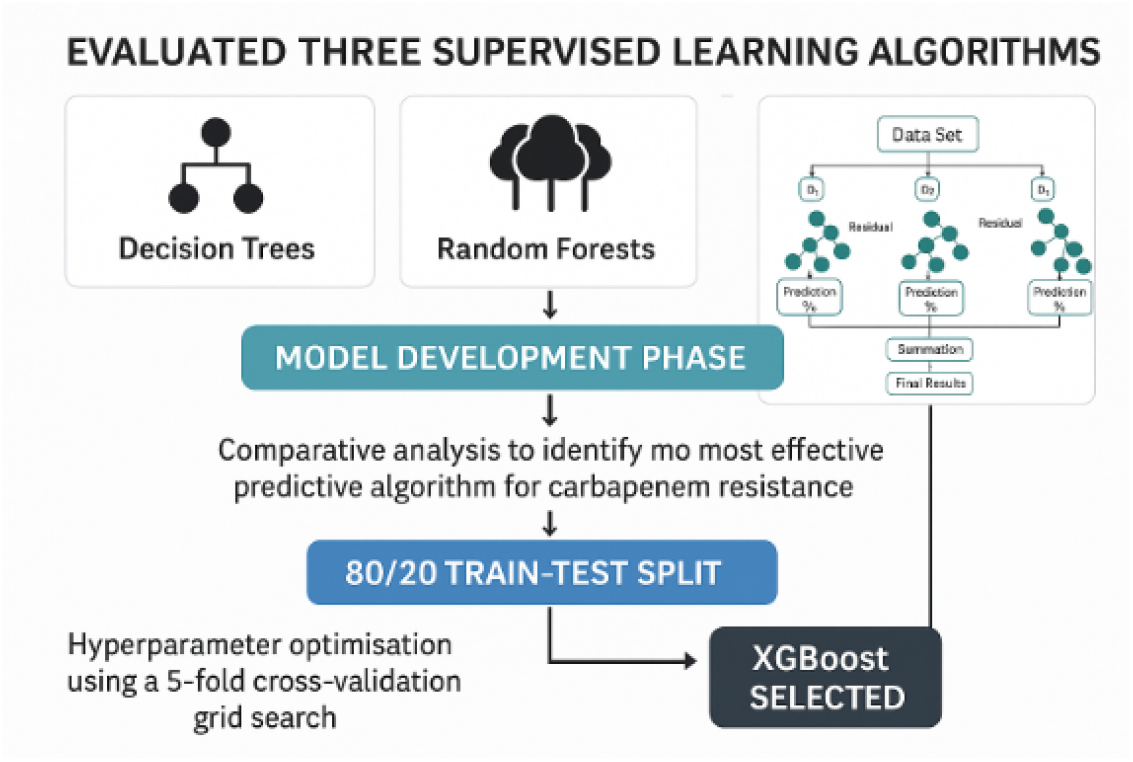
Process of model selection.

**Image 3.**
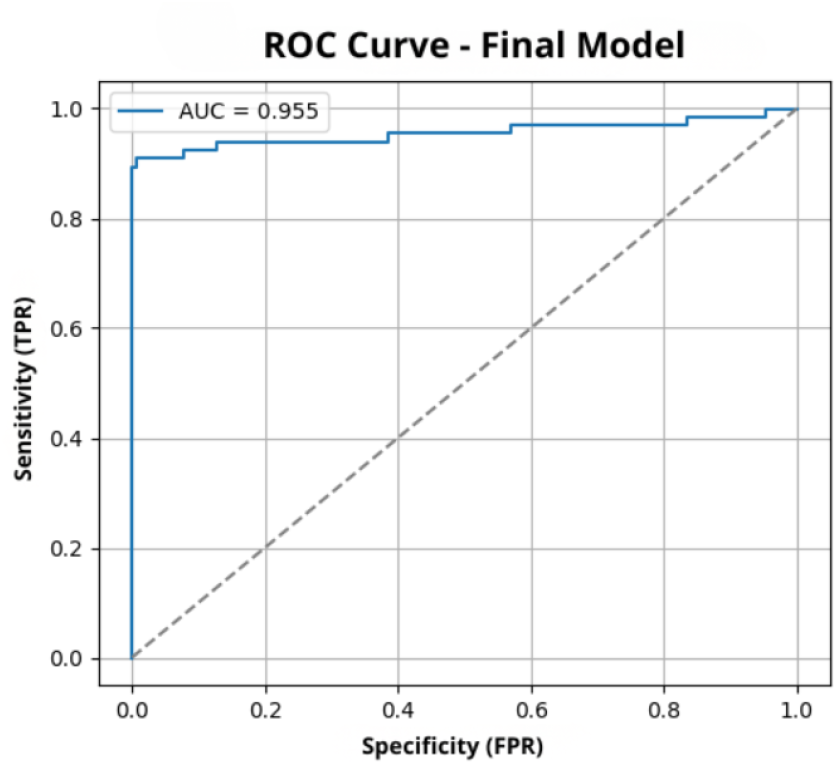
AUC-ROC curve for our XGBoost model.

**Image 4.**
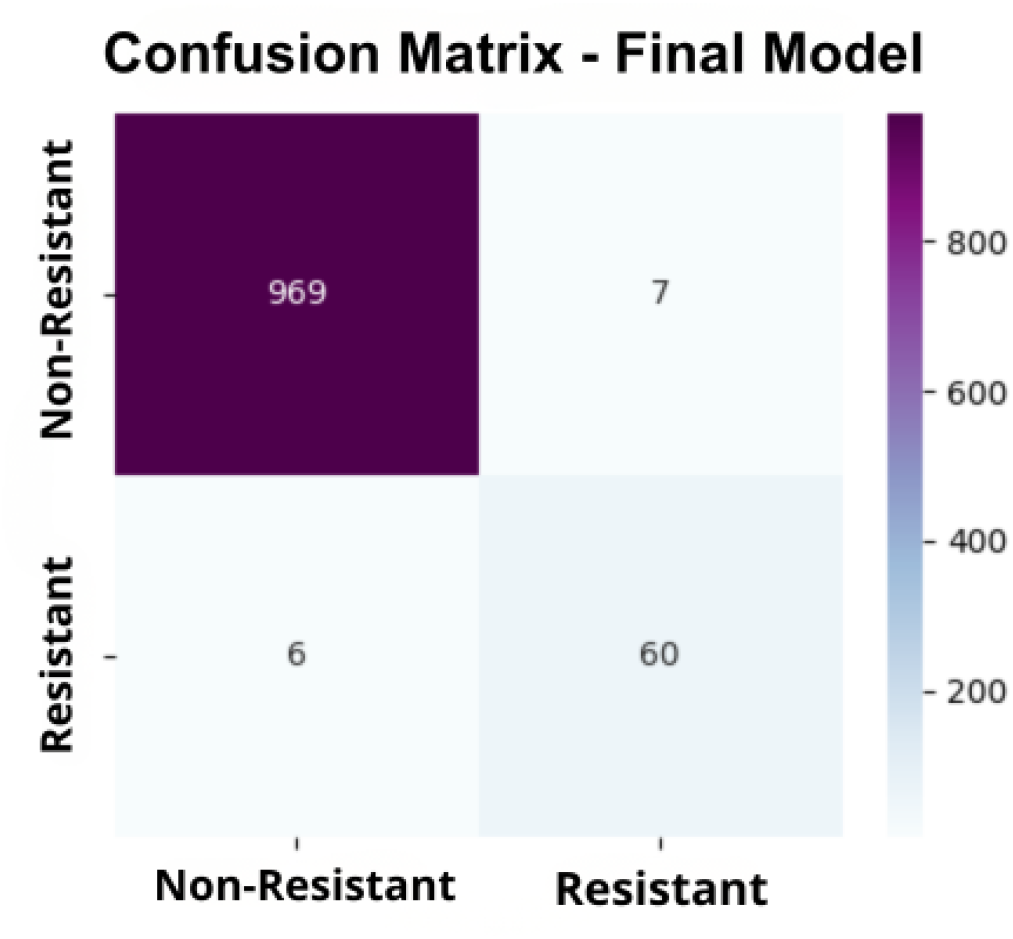
Confusion Matrix Results. Performance of the Carbapenem Resistance Predictive Model. This model achieved excellent discriminative ability. It showed high sensitivity (90.9%) and specificity (99.3%), demonstrating its clinical usefulness in anticipating carbapenem resistance.

**Image 5.**
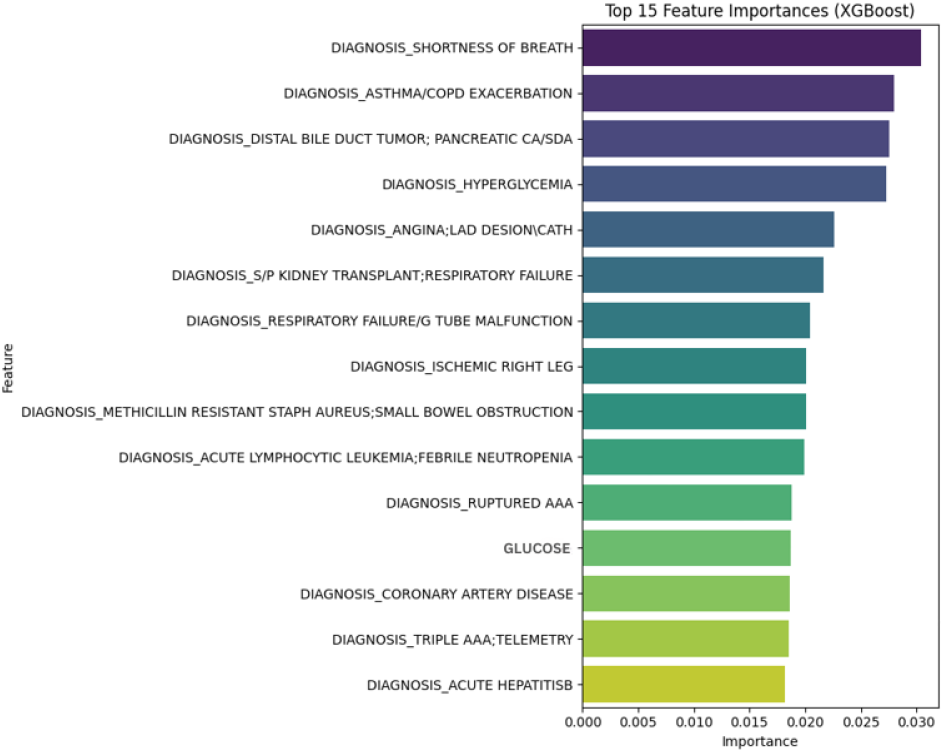
Feature Importance.

### Feature Selection and Data Preprocessing

The data extracted included:

- **Demographics:** age, sex, ethnicity
- **Clinical factors:** vital signs, ICU admission, severity of illness
- **Comorbidities:** diabetes, chronic kidney disease, malignancy, cardiovascular disease and more
- **Antibiotic history:** prior exposure, previous antibiotic administered
- **Laboratory biomarkers:** White Blood Cell count, hemoglobin, creatinine, lactate, CRP protein
- **Microbiological-related variables:** microorganism identification, culture source, susceptibility pattern
- **Admission factors:** emergency vs. elective, hospital service, length of stay

### Outcome Definition

The primary outcome for this predictive model is the presence of carbapenem resistance. This crucial predicted variable, labelled ‘RESISTANT’ in the database, used as the prediction (y) in the computer model, was meticulously derived from the ‘MICROBIOLOGYEVENTS’ dataset. Patients were classified as ‘resistant’ if at least one microbiology event indicated a positive culture with an INTERPRETATION of ‘R’ (resistant) to any of the specified carbapenem antibiotics, mainly meropenem, imipenem, or ertapenem. This approach enabled the creation of a clear binary target variable that distinguished between patients with carbapenem resistance and those without, thereby providing the basis for the model’s classification task.

### Preprocessing

We performed meticulous data preparation for the carbapenem resistance prediction model. The process involved several key steps: initially, diagnostics, admission date, and numerical clinical and laboratory features such as age, ICU admission status, time to ICU entry, and various blood test results (blood tests, leukocytes, hemoglobin, creatinine, glucose, and C-Reactive Protein (CRP)) were selected. Missing values meant a discarded data point in our model. Concurrently, categorical demographic and admission-related variables, including primary diagnosis, admission type, admission location, insurance, ethnicity, and marital status, were scrutinized during hyperparameter optimization of our modelling state. The ‘DIAGNOSIS’ variable’s high cardinality was addressed by extracting its initial component, then encoding all categorical features with a flag set to ‘drop_first=True’ to convert them to numerical values while avoiding multicollinearity. Furthermore, a binary flag, ‘HAD_ANTIBIOTIC’, was included from the ‘MICROBIOLOGY EVENTS’ dataset to indicate prior antibiotic exposure, recognizing its significance in resistance prediction. Finally, all processed numerical and encoded categorical features, along with the antibiotic exposure flag, were concatenated to construct the comprehensive input dataset ‘x’, with ‘RESISTANT’ (carbapenem resistance) as the binary target variable.

These comprehensive preprocessing steps were essential for transforming raw, heterogeneous data into a structured and analyzable format suitable for machine learning. Careful handling of missing values, encoding of categorical variables, and engineering of relevant features such as prior antibiotic exposure optimized the dataset ‘x’ to provide the model with informative inputs. This preparation minimized noise, addressed potential biases, and ensured that the predictive model for carbapenem resistance was built on a robust foundation, thereby enhancing its ability to learn meaningful patterns and generalize effectively to unseen patient data.

### Modeling

We evaluated three supervised learning algorithms:

1. **Decision Trees**
2. **Random Forests**
3. **Extreme Gradient Boosting (XGBoost)**

The model development phase included a rigorous comparative analysis to identify the most effective predictive algorithm for carbapenem resistance. This systematic approach, utilizing multiple supervised learning techniques, ensured that the selected model was robust and generalizable. Initially, an 80/20 train-test split created a distinct set for unbiased evaluation, preventing memorization of the training data. Hyperparameter optimization using a 5-fold cross-validation grid search was applied to all candidate models to fine-tune performance and mitigate overfitting, resulting in the selection of XGBoost for its superior predictive performance. This selection process aligns with best practices in machine learning, ensuring a reliable and high-performing solution for clinical prediction.

### Code Availability

The complete source code and accompanying resources for this project are publicly available on GitHub. You can access the repository at: https://github.com/SDCUCS/ML-Carbapenemic-Resistance-MIMIC-III

### Variables Impacts

Assessing the impact of individual variables on the model’s predictions is essential for clinical interpretability and for identifying key factors associated with carbapenem resistance. This section elucidates which clinical, demographic, and laboratory features most significantly influence the model’s decision-making process. Techniques such as feature importance from tree-based models (XGBoost being the best performing). This analysis pinpoints the most influential predictors, providing insights into the mechanisms of resistance and informing future clinical interventions or research.

## Results

### Model Evaluation, Evaluation metrics proposed

Key performance metrics included:

- AUC-ROC
- Precision
- Sensitivity
- Specificity
- Confusion matrix analysis

Metrics were computed on the testing dataset to prevent optimistic bias.

### Model Performance

Among the evaluated algorithms, Extreme Gradient Boosting (XGBoost) demonstrated superior predictive performance for carbapenem resistance, achieving an AUC of 0.955. Detailed analysis revealed a precision of 0.989, sensitivity of 0.909, and specificity of 0.993. These robust metrics, particularly the high true-negative and true-positive rates observed in the confusion matrix and a minimal false-positive rate, confirm the model’s ability to accurately discriminate between resistant and non-resistant infections. These characteristics highlight their potential for clinical implementation as a reliable early detection tool. Interpretability analysis identified several critical predictors that align with established clinical mechanisms of antibiotic resistance, including prior carbapenem exposure, ICU admission at the time of culture, elevated lactate levels, abnormal creatinine levels, white blood cell count abnormalities, recent severe bacterial infection, and prior hospitalization within 90 days. These insights enhance the model’s credibility and provide valuable guidance for clinical decision-making.

XGBoost demonstrated the highest predictive performance:

- **AUC:** 0.955
- **Precision:** 0.989
- **Sensitivity:** 0.909
- **Specificity:** 0.993

These results indicate that the model accurately discriminates between resistant and non-resistant infections with minimal false positives.

### Confusion Matrix Interpretation

The confusion matrix revealed:

- High true-negative rate
- High true-positive rate
- Low false-positive rate
- Incorporation of multiple clinically relevant variables

These characteristics make the model a strong candidate for clinical implementation.

### Feature Importance

Additionally, a diagnosis of recent severe bacterial infection and prior hospitalization within 90 days emerged as critical predictive factors. These findings are consistent with established biological and clinical mechanisms underlying antibiotic resistance, providing valuable insights into the multifactorial nature of this challenge and reinforcing the model’s clinical relevance.

The permutation feature importance analysis of the Extreme Gradient Boosting (XGBoost) model revealed critical predictors of carbapenem resistance, albeit with nuances that diverged from initial clinical hypotheses. The most prominent features were largely dominated by highly specific diagnostic categories, such as ‘DIAGNOSIS_SHORTNESS OF BREATH’, ‘DIAGNOSIS_ASTHMA/COPD EXACERBATION’, and ‘DIAGNOSIS_HYPERGLYCEMIA’, among others. This suggests that the model leveraged granular diagnostic information as primary indicators of resistance within the dataset. Notably, among the general biochemical markers, only ‘GLUCOSE’ (glucose levels) ranked within the top 15 most important features. Conversely, several clinically anticipated predictors, including ‘previous carbapenem exposure’, ‘ICU admission at culture time’, and other key laboratory values such as lactate and creatinine, did not appear among the highest-ranking features. This discrepancy highlights the data-driven nature of the model’s insights, emphasizing the predictive power of detailed diagnostic contexts captured in the dataset, and warrants further investigation into the interplay between these diagnoses and carbapenem resistance mechanisms.

## Discussion

This work demonstrates the viability and reliability of applying machine learning techniques to predict carbapenem resistance using routinely collected clinical data. The high discriminative performance of XGBoost indicates that machine learning models can integrate diverse predictors and identify at-risk patients earlier than standard clinical approaches. Early prediction is essential, as delays in effective therapy are strongly associated with increased mortality in patients with resistant infections. Conversely, unnecessary carbapenem exposure accelerates the spread of resistance. Therefore, accurate predictive tools may improve both individual patient outcomes and institutional antibiotic stewardship.

Previous studies on antimicrobial resistance prediction have often been limited by small sample sizes, narrow clinical settings, or a lack of external validation (Pinto de Moura et al., 2022). In contrast, our dataset includes a large number of patients, a diverse range of clinical variables, and robust methodological steps to minimize bias.

## Limitations

Despite its robust performance, this study has several limitations that warrant consideration. The retrospective design, while valuable for initial exploration, introduces potential biases and dependencies on the quality of historical data. Additionally, the model may be susceptible to unmeasured confounders not captured within the available dataset, which could influence the observed associations. To approach and address these limitations and enhance the model’s generalizability and predictive accuracy, future research should prioritize multi-center external validation. Integrating advanced genetic resistance markers could also provide a more comprehensive understanding of antibiotic resistance mechanisms, further refining predictive capabilities and supporting more precise clinical interventions. Intelligence and programming in cloud-based infrastructure platforms offer a promising avenue for enhancing early detection of carbapenem resistance in hospitalized patients. By integrating multidimensional, multivariate, clinical, laboratory and microbiological data, the XGBoost model developed in this study demonstrates strong predictive accuracy and practical utility for antimicrobial stewardship programs. Real-time implementation in hospital systems may improve clinical outcomes, reduce inappropriate antibiotic use, and contribute to global efforts to combat antimicrobial resistance. Moreover, we could also update the study reported here using the newly released MIMIC-IV database.

## Data Availability

Data Availability Statement: The data that support the findings of this study are available in the MIMIC-III Clinical Database (version 1.4), hosted on the PhysioNet repository. Access to the data is restricted and requires successful completion of the mandated human subjects research training (e.g., CITI training) and signing a Data Use Agreement. The MIMIC-III Clinical Database is available here: https://doi.org/10.13026/C2XW26 (PhysioNet). All code and derived data used for this analysis are available from the corresponding author upon reasonable request.

